# Prevention of COVID-19 by mRNA-based vaccines within the general population of California

**DOI:** 10.1101/2021.04.08.21255135

**Authors:** Kristin L. Andrejko, Jake Pry, Jennifer F. Myers, Nicholas P. Jewell, John Openshaw, James Watt, Seema Jain, Joseph A. Lewnard, on behalf of the California COVID-19 Case-Control Study Team

## Abstract

**Background:** Estimates of COVID-19 vaccine effectiveness under real-world conditions, and understanding of barriers to uptake, are necessary to inform vaccine rollout.

**Methods:** We enrolled cases (testing positive) and controls (testing negative) from among the population whose SARS-CoV-2 molecular diagnostic test results from 24 February-29 April 2021 were reported to the California Department of Public Health. Participants were matched on age, sex, and geographic region. We assessed participants’ self-reported history of COVID-19 vaccine receipt (BNT162b2 and mRNA-1273). Participants were considered fully vaccinated two weeks after second dose receipt. Among unvaccinated participants, we assessed willingness to receive vaccination, when eligible. We measured vaccine effectiveness (VE) via the matched odds ratio of prior vaccination, comparing cases with controls.

**Results:** We enrolled 1023 eligible participants aged ≥18 years. Among 525 cases, 71 (13.5%) received BNT162b2 or mRNA-1273; 20 (3.8%) were fully vaccinated with either product. Among 498 controls, 185 (37.1%) received BNT162b2 or mRNA-1273; 86 (16.3%) were fully vaccinated with either product. Two weeks after second dose receipt, VE was 86.8% (95% confidence interval: 68.6-94.7%) and 85.6% (69.1-93.9%) for BNT162b2 and mRNA-1273, respectively. Fully vaccinated participants receiving either product experienced 91.3% (79.7-96.3%) and 68.3% (28.5-86.0%) VE against symptomatic and asymptomatic infection, respectively. Among unvaccinated participants, 42.4% (159/375) residing in rural regions and 23.8% (67/281) residing in urban regions reported hesitancy to receive COVID-19 vaccination.

**Conclusions:** Authorized mRNA vaccines are effective at reducing documented SARS-CoV-2 infections within the general population of California. Vaccine hesitancy presents a barrier to reaching coverage levels needed for herd immunity.

**Brief points:**

- Vaccination is preventing documented SARS-CoV-2 infection in California, with 68% and 91% effectiveness against asymptomatic and symptomatic infection, respectively.
- Vaccine effectiveness was equivalent for BNT126b2 and mRNA-1273.
- Only 66% of unvaccinated participants were willing to receive the vaccine when eligible.

## INTRODUCTION

After being found safe and efficacious in preventing coronavirus disease 2019 (COVID-19) in randomized controlled trials [1–3], vaccines against severe acute respiratory syndrome coronavirus 2 (SARS-CoV-2) are now being administered to the general public under emergency use authorization. Two mRNA-based vaccines encoding the SARS-CoV-2 spike protein, BNT162b2 (Pfizer/BioNTech) and mRNA-1273 (Moderna), have been the main products in use since December 2020. By early May, 2021, 40% of California residents were considered fully vaccinated [4].

Observational studies characterizing COVID-19 vaccine effectiveness (VE) are needed to understand performance under real-world conditions [5], for instance evaluating VE against clinical endpoints not addressed in trials, monitoring VE as novel SARS-CoV-2 variants emerge and circulate, and defining VE for dosing schedules that differ from those assessed in trials [6]. While most studies of real-world VE to date have followed healthcare workers and other essential or frontline personnel [7–9], vaccine eligibility has rapidly expanded to included broader population groups over the winter and spring of 2021 throughout the United States. In California, vaccination was made available to healthcare workers December 14, 2020, and expanded to include persons at increased risk due to older age or occupation (including workers in emergency services, food and agriculture, or childcare and education) during January and February, 2021. Eligibility was extended to persons aged 16-64 years with high-risk medical conditions in March, 2021, and has included all persons aged ≥16 years since April 15, 2021. To inform ongoing vaccination efforts, it is thus crucial to understand VE within the general population, and to identify reasons behind individuals’ decisions to delay or defer vaccination.

In conjunction with epidemiologic surveillance, we initiated a test-negative case-control study design to monitor VE within the general population of California in real time. Over the study period summarized here (February 24, 2021 to April 29, 2021), sequenced SARS-CoV-2 isolates in California were predominantly identified as B.1.427/429 (50-60%) variants in February and March; by April, B.1.1.7 variants overtook other lineages and accounted for 49% of sequenced SARS-CoV-2 isolates, as compared to 6% in February, while the proportion of B.1.427/429 variants declined to ∼20% [10]. Here we provide an early assessment of VE for authorized mRNA-based COVID-19 vaccines, and report data on the intentions of unvaccinated participants to receive vaccination.

## METHODS

### Design

All diagnostic tests in California for SARS-CoV-2 are reported by laboratories and medical providers to their local health jurisdiction (LHJ). Sixty of 61 LHJs report data directly to the California Department of Public Health (CDPH) via a web-based reporting system, while Los Angeles County transmits data daily via an electronic file. California residents with molecular SARS-CoV-2 test results (e.g., polymerase chain reaction [PCR]) between 24 February-April 29, 2021 and a telephone number were eligible for participation in this study. Cases were defined as persons with positive molecular SARS-CoV-2 test results during the study timeframe, whereas controls were persons with negative SARS-CoV-2 molecular test results during the same period.

Each day during the study period, we prospectively selected cases with a telephone number and newly-reported positive molecular test result within each of nine regions of the state, sampling cases at random with intent to enroll equally across each region of the state (**Table S1**). For each case who consented and completed the study interview, we attempted to enroll and interview one control from a sample of 30 controls randomly selected to match the case by age group (18-39, 40-64, ≥65 years), sex, region, and week of SARS-CoV-2 test. A maximum of two call attempts were made for each case and control. Call shifts were scheduled to cover morning, afternoon, and evening periods of each day.

To mitigate bias potentially resulting from previous infection-derived immunity [6], participants who recalled receiving any previous positive test result for SARS-CoV-2 infection or seropositivity, prior to the reported test, were not eligible to continue the interview. This analysis excludes data from children aged 0-17 years, who were generally ineligible for COVID-19 vaccination over the study period; and participants who reported receiving COVID-19 vaccinations other than BNT162b2 or mRNA-1273 (due to limited coverage of a third authorized vaccine, Ad26.COV2.S, over the study period), or receipt of COVID-19 vaccination but without knowledge of precise dates of vaccination.

### Exposures

We designed and implemented a standardized questionnaire to be delivered via facilitated telephone interviews in English or Spanish collecting data on participant demographics, symptoms, and vaccination status. We asked participants to indicate whether they had received any COVID-19 vaccine, and to reference their COVID-19 vaccination card to report the manufacturer, number, and dates of doses received. We also asked unvaccinated participants whether they would be willing to receive a COVID-19 vaccine when available to them; if participants indicated they were not likely to receive a vaccine or unsure, we asked for participants to state any and all reasons behind their hesitancy. Additionally, we asked participants to indicate the reason they sought a COVID-19 test, and presence of any COVID-19 symptoms within the 14 days prior to their test date (**Supplementary File S1)**.

The study protocol was granted a non-research determination by the State of California Health and Human Services Agency Committee for the Protection of Human Subjects (project number: 2021-034).

### Statistical analysis

Our primary study objective was to estimate VE of two doses of BNT162b2 or mRNA-1273 against documented SARS-CoV-2 infection, two weeks after receipt of the second dose of either vaccine. To estimate VE under the test-negative study design, we calculated the Mantel Haenszel (matched) odds ratio (OR_MH_) of vaccination among cases relative to controls [5,6]. We used conditional logistic regression models defining match strata by age group, sex, region, and testing week to estimate the OR_MH_ (and accompanying 95% confidence interval [CI]). We defined vaccine exposures as receipt of two doses of BNT162b2 or mRNA-1273 at least two weeks before participants’ date of testing; unvaccinated status was considered the reference exposure. We calculated adjusted VE as (1–OR_MH_)×100%. We determined that interim analyses with 500 cases and 500 controls enrolled would provide 90% statistical power for estimating VE of 55% or greater at the two-sided *p*<0.05 confidence threshold, assuming 10% of controls would be fully vaccinated. We did analyses in R software (version 3.6.1; R Foundation for Statistical Computing; Vienna, Austria).

As secondary analyses, we also aimed to assess VE for incomplete vaccination series, VE for each product, and VE against SARS-CoV-2 infection endpoints corresponding to differing levels of clinical severity. To determine VE for incomplete vaccination series, we defined exposures as receipt of 1 dose or 2 doses of BNT162b2 or mRNA-1273 within 1-7 or 8-14 days before participants’ date of testing, or 1 dose of BNT162b2 or mRNA-1273 ≥15 days before participants’ date of testing. As described above, we used conditional logistic regression models to compute the OR_MH_ among cases relative to controls.

To determine product-specific VE, we restricted the vaccinated population to participants who received two doses of either BNT162b2 or mRNA-1273 ≥15 days before their date of testing. To determine VE against differing clinical endpoints, we conducted analyses restricting cases to participants testing positive with symptoms; without symptoms; who were hospitalized for COVID-19; who reported seeking healthcare or advice via outpatient or virtual interactions with healthcare providers; and who did not seek treatment or advice from a healthcare provider beyond receipt of a molecular SARS-CoV-2 diagnostic testing. Each of these groupings of cases was compared against match-eligible controls to compute the OR_MH_ of vaccination (defined as two doses received ≥15 days prior, versus no doses received), using the same conditional logistic regression framework described above. For these secondary analyses, we compared fully vaccinated and unvaccinated participants only, as sufficient counts were not available to stratify VE estimates by doses received and time since receipt.

Last, to understand factors predicting vaccine hesitancy among participants who had not yet received any doses of a COVID-19 vaccine, we fit logistic regression models defining hesitancy to receive vaccination as the outcome; covariates selected *a priori* for inclusion as potential causal factors were age group, region, sex, income, and race/ethnicity. Participants who reported being unwilling or unsure about receiving a COVID-19 vaccine when eligible were considered vaccine-hesitant. As missing data were present in participants’ responses regarding income (189/656; 28.8%) and race (10/656; 1.5%), we conducted analyses of vaccine hesitancy across five datasets generated through multiple imputation by chained equations using the Amelia II package in R [11]. Under the assumption that data were missing conditionally at random, given observations of other covariates, all variables included in the analyses model were included in the imputation models. We compared measures of association to those resulting from complete-case analysis without imputation as a supplemental check.

## RESULTS

From February 24 to April 29, 2021, there were 4,827,165 SARS-CoV-2 molecular test results reported to CDPH with a telephone number and identification of age, sex, and region of participant (108,606 positive and 4,718,559 negative; **Figure 1**). Calls were placed to 3847 cases and 5253 controls, among whom we enrolled 603 cases (15.7%) and 590 controls (11.2%). Among participants enrolled, 78 cases and 92 controls who were ineligible for the analyses reported here, including participants who were <18 years old, received COVID-19 vaccines other than BNT162b2 or mRNA-1273, or were unable to provide precise dates of COVID-19 vaccine receipt. Our final study population for the primary analysis included 525 cases and 498 controls (**Table 1**; **Table S2**). Among participants enrolled, 20.9% (214/1023) were contacted within ≤3 days of their test results being posted, and 98.3% (1006/1023) were contacted within ≤7 days of their test results being posted.

**Figure 1:**
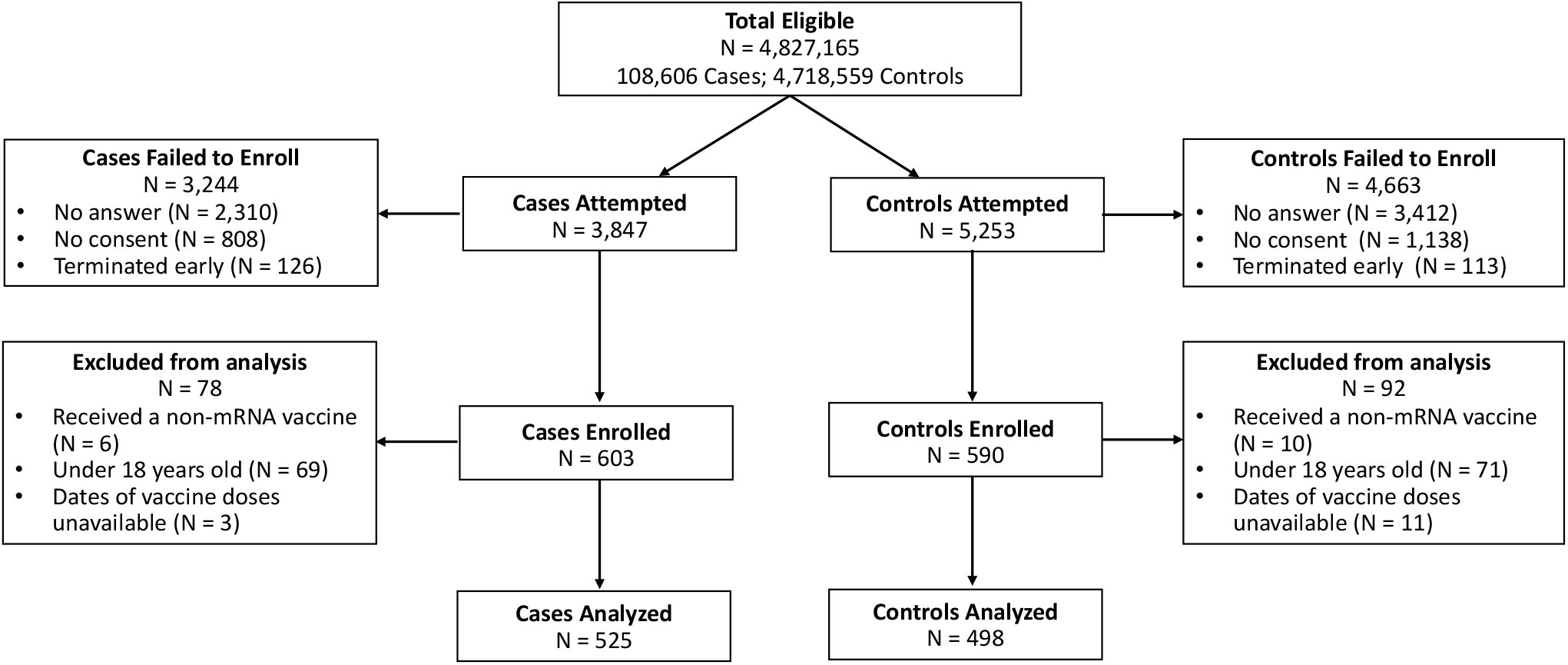
Enrollment of participants in the California COVID-19 Case-Control study. Data in the figure indicate numbers of tests reported, cases and controls for whom contact was attempted, and excluded and enrolled participants for this analysis.

**Table 1:** Distribution of cases and controls.

|  |  | Overall<br>*n* (%)<br>*N* = 1023 | Case<br>*n* (%)<br>*N* = 525 | Control<br>*n* (%)<br>*N* = 498 |
| --- | --- | --- | --- | --- |
| Age | 18-29 | 395 (38.6) | 200 (38.1) | 195 (39.2) |
|  | 30-49 | 363 (35.5) | 188 (35.8) | 175 (35.1) |
|  | 50-64 | 192 (18.8) | 100 (19.0) | 92 (18.5) |
|  | 65+ | 73 ( 7.1) | 37 ( 7.0) | 36 ( 7.2) |
| Region | *Predominantly urban regions* |  |  |  |
|  | San Francisco Bay Area | 129 (12.6) | 66 (12.6) | 63 (12.7) |
|  | Greater Los Angeles Area | 91 ( 8.9) | 48 ( 9.1) | 43 ( 8.6) |
|  | Greater Sacramento Area | 115 (11.2) | 58 (11.0) | 57 (11.4) |
|  | San Diego and southern Border | 110 (10.8) | 54 (10.3) | 56 (11.2) |
|  | *Predominantly rural regions* |  |  |  |
|  | Central Coast | 140 (13.7) | 74 (14.1) | 66 (13.3) |
|  | Northern Sacramento Valley | 116 (11.3) | 60 (11.4) | 56 (11.2) |
|  | San Joaquin Valley | 106 (10.4) | 54 (10.3) | 52 (10.4) |
|  | Northwestern California | 108 (10.6) | 55 (10.5) | 53 (10.6) |
|  | Sierras Region | 108 (10.6) | 56 (10.7) | 52 (10.4) |
| Sex | Male | 519 (50.7) | 264 (50.3) | 255 (51.2) |
|  | Female | 504 (49.3) | 261 (49.7) | 243 (48.8) |
| Household income | Under \$50,000 | 272 (26.6) | 153 (29.1) | 119 (23.9) |
| | \$50,000 to \$100,000 | 220 (21.5) | 113 (21.5) | 107 (21.5) |
| | \$100,000 to \$150,000 | 121 (11.8) | 45 ( 8.6) | 76 (15.3) |
| | Over \$150,000 | 135 (13.2) | 64 (12.2) | 71 (14.3) |
|  | Refuse | 154 (15.1) | 86 (16.4) | 68 (13.7) |
|  | Not sure | 121 (11.8) | 64 (12.2) | 57 (11.4) |
| Race/Ethnicity | White | 444 (43.4) | 217 (41.4) | 227 (45.6) |
|  | Hispanic | 286 (28.0) | 160 (30.5) | 126 (25.3) |
|  | Asian | 115 (11.3) | 58 (11.1) | 57 (11.4) |
|  | Black | 47 ( 4.6) | 30 ( 5.7) | 17 ( 3.4) |
|  | More than 1 race | 89 ( 8.7) | 36 ( 6.9) | 53 (10.6) |
|  | Native American | 16 ( 1.6) | 11 ( 2.1) | 5 ( 1.0) |
|  | Native Hawaiian | 10 ( 1.0) | 4 ( 0.8) | 6 ( 1.2) |
|  | Refuse | 15 ( 1.5) | 8 ( 1.5) | 7 ( 1.4) |
| Vaccination | Unvaccinated | 767 (75.0) | 454 (86.5) | 313 (62.9) |
|  | Incompletely vaccinated | 150 (14.7) | 51 ( 9.7) | 99 (19.9) |
|  | Fully vaccinated<sup>1</sup> | 106 (10.4) | 20 ( 3.8) | 86 (17.3) |

<sup>1</sup>An individual was considered "fully-vaccinated" ≥ 14 days after two doses of Pfizer/BioNTech BNT162b2 or Moderna mRNA-1273, and "incompletely-vaccinated" if they received only one dose or two doses <14 days after second dose

Among 525 cases, 288 (54.9%) indicated they were tested due to concerns about symptoms. Of these 288 symptomatic cases, 262 (91.0%) were unvaccinated and 26 (9.0%) received ≥1 vaccine dose (**Table 2**). In contrast, among 498 controls, 56 (11.2%) indicated seeking testing due to symptoms, among whom 43 (76.8%) were unvaccinated and 13 (23.2%) received ≥1 vaccine dose. The most commonly indicated reason for testing among controls was routine screening required for work or school attendance (233/498; 46.8%), whereas the most common reasons for testing among cases were symptoms (288/525; 54.9%) and known contact with a positive case (173/525; 33.0%).

**Table 2:** Reasons for testing.

| Reasons* | Controls |  | Cases |  |
| --- | --- | --- | --- | --- |
|  | Unvaccinated | Vaccinated<sup>1</sup> | Unvaccinated | Vaccinated |
|  | N=313 | N=185 | N=454 | N=71 |
| Contact with positive case | 28 (8.9) | 8 (4.3) | 143 (31.5) | 30 (42.3) |
| Contact with symptomatic individual | 12 (3.8) | 4 (2.2) | 18 (4.0) | 2 (2.8) |
| Told by public health worker to get tested | 1 (0.3) | 1 (0.5) | 3 (0.7) | 0 (0.0) |
| Routine screening for my work or school | 120 (38.3) | 113 (61.1) | 29 (6.4) | 17 (23.9) |
| Test required for medical procedure or hospital admittance | 43 (13.7) | 25 (13.5) | 16 (3.5) | 5 (7.0) |
| Someone in household had contact with a positive case | 4 (1.3) | 0 (0.0) | 11 (2.4) | 0 (0.0) |
| Test required to attend public event/ share public space | 2 (0.3) | 0 (0.0) | 1 (0.5) | 0 (0.0) |
| I just wanted to see if I was infected | 71 (22.7) | 18 (9.7) | 43 (9.5) | 4 (5.6) |
| Concerned about symptoms | 43 (13.7) | 13 (7.0) | 262 (57.7) | 26 (36.6) |
| Pre or post-travel screening | 21 (6.7) | 7 (3.8) | 17 (3.7) | 4 (5.6) |

\*Since interviewers indicated all reasons listed by participants, reasons will not sum to the total sample size.
<sup>1</sup>An individual is considered vaccinated if they have had at least one dose of a SARS-CoV-2 mRNA vaccine.

Among 525 cases, 43 (8.2%) and 28 (5.3%) reported receiving at least one dose of BNT162b2 and mRNA-1273, respectively (**Figure 2**; **Table 1**; **Table S3**). Among 498 controls, 98 (19.7%) and 87 (17.5%) received at least one dose of BNT162b2 and mRNA-1273, respectively. Twenty cases (3.8% of 525) and 86 (17.3% of 498) controls were fully vaccinated with either product, with ≥15 days passing from receipt of their second dose to the date of testing. For fully-vaccinated participants receiving either BNT162b2 or mRNA-1273, VE was 87.4% (95%CI: 77.1-93.1%).

**Figure 2:**
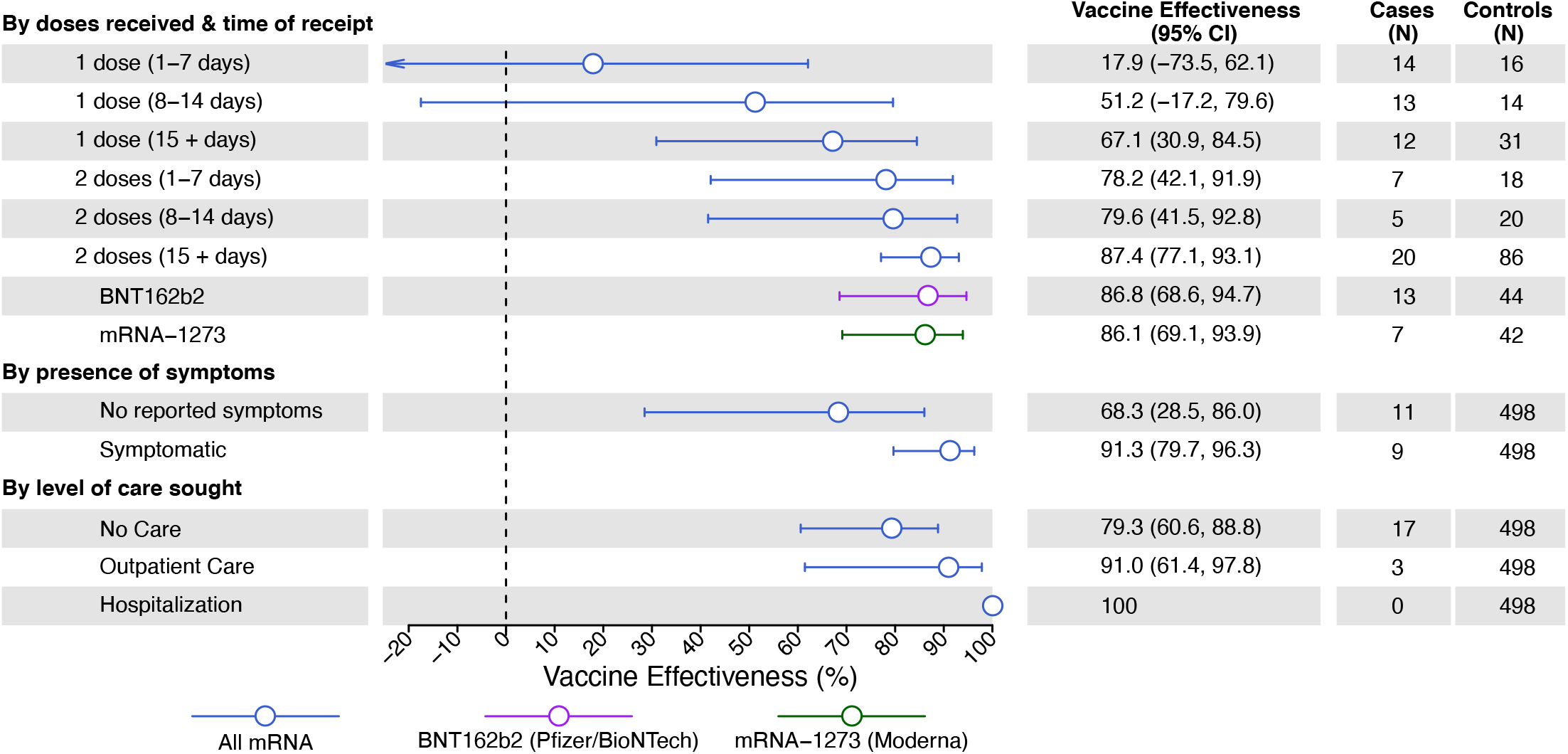
COVID-19 vaccine effectiveness, by doses received and time since last dose. Lines denote 95% confidence intervals, respectively, for estimates of vaccine effectiveness. Estimates were calculated via conditional logistic regression.

We did not identify protection within the first 7 days after receipt of a first BNT162b or mRNA-1273 dose (VE: 17.9%; 95%CI: –73.5-62.1%). Within the second week after receipt of a first dose for either vaccine, VE was 51.2% (95%CI: –17.2-79.6%); ≥15 days after receipt of a first dose, and before receipt of a second dose, VE was 67.1% (95%CI: 30.9-84.5%). Following receipt of a second dose, VE was 78.2% (95%CI: 42.1-91.9%) at days 1-7, 79.6% (95%CI: 41.5-92.8%) at days 8-14. VE estimates were similar in analyses that restricted or did not restrict the sample to participants who reported consulting their vaccination cards or calendars during the telephone interview to confirm dates of receipt of each dose (**Figure S1**).

Estimated protection among fully-vaccinated participants did not differ according to the product received; among recipients of BNT162b and mRNA-1273, VE was 86.8% (95%CI: 68.6-94.7%) and 86.1% (95%CI: 69.1-93.9%), respectively (**Figure 2**).

Among fully vaccinated cases, 45.0% (9/20) reported at least one symptom, in contrast to 78.0% (354/454) of unvaccinated cases, 66.7% (34/41) of partially vaccinated cases, and 13.7% (68/498) of controls (**Table S4**). For symptomatic and asymptomatic infection endpoints, VE was 91.3% (95%CI: 79.7-96.3%) and 68.3% (95%CI: 28.5-86.0%), respectively, at ≥15 days after the second dose (**Figure 2**). Eighteen (3.4%) of 525 cases were hospitalized by the time of our telephone interview, among whom 15 (83.3%) were unvaccinated, and three (16.7%) were partially vaccinated (**Table S4**). Among all 525 cases, 150 (28.6%) sought treatment, care, or advice via outpatient or virtual interactions with healthcare providers, among whom 132 (25.1%) were unvaccinated, 15 (2.9%) were incompletely vaccinated, and 3 (0.6%) were fully vaccinated. Among 128 cases who did not experience symptoms, 103 (80.4%) did not seek care. Considering these differing levels of care sought for SARS-CoV-2 infection, VE was 79.3% (95%CI: 60.6-88.8%) against episodes for which cases did not seek treatment or advice, 91.0% (95%CI: 61.4-97.8%) against episodes for which cases sought healthcare through outpatient or virtual interactions, and 100% (with undefined confidence limits) against hospitalized episodes (**Figure 2**).

Overall, 226 (34.5%) of 656 unvaccinated participants (including 139/403 [34.5%] unvaccinated cases and 87/253 [34.4%] unvaccinated controls) indicated they were unlikely or unsure about receiving COVID-19 vaccination when eligible (**Tables 3; Table S5; Table S6)**. Residents of rural regions had 2.42 (95%CI 1.66-3.52) higher adjusted odds of reporting they were unlikely or unsure about receiving vaccination, when eligible, whereas hesitancy to receive vaccination was not independently associated with age or household income. Adjusted odds of reporting hesitancy to receive vaccination were 1.47 (95%CI 1.04-2.08) higher among females compared with males. In comparisons by participants’ race/ethnicity, adjusted odds of reporting hesitancy to receive vaccination were 2.54 (95%CI 1.24-5.15) higher among non-Hispanic Black participants than non-Hispanic Whites; in contrast, adjusted odds of vaccine hesitancy were 0.72 (95%CI: 0.46-1.12) fold as high among Hispanic participants as among non-Hispanic whites. Point estimates of odds ratios were similar in complete-case analyses without imputation (**Table S7**). Fears over vaccine side effects (66/219 [30.1%]) or safety (60/219 [27.4%]) were the most common concerns among participants expressing hesitancy to receive vaccination (**Table 4**).

**Table 3:** Predictors of vaccine hesitancy.

| Participant characteristics | Enthusiasm to receive vaccination |  | Odds ratio (95% CI) |  |
| --- | --- | --- | --- | --- |
|  | Not willing/unsure, n (%)<br>N=226 | Willing, n (%)<br>N=430 | Unadjusted | Adjusted |
| Case status<sup>1</sup> |  |  |  |  |
| Case with SARS-CoV-2 infection | 139 (61.5) | 264 (61.4) | N/A | N/A |
| Uninfected control | 87 (38.5) | 166 (38.6) | N/A | N/A |
| Age |  |  |  |  |
| 18-29 | 82 (36.3) | 189 (44.0) | Ref. | Ref. |
| 30-49 | 93 (41.2) | 147 (34.2) | 1.45 (1.01,2.10) | 1.45 (0.97,2.16) |
| 50-64 | 34 (15.0) | 76 (17.7) | 1.03 (0.64,1.66) | 0.77 (0.46,1.28) |
| 65+ | 17 (7.5) | 18 (4.2) | 2.20 (1.07,4.40) | 1.66 (0.77,3.57) |
| Region |  |  |  |  |
| Predominantly urban regions<sup>2</sup> | 67 (29.6) | 214 (49.8) | Ref. | Ref. |
| Predominantly rural regions<sup>3</sup> | 159 (70.4) | 216 (50.2) | 2.35 (1.66,3.29) | 2.42 (1.66,3.52) |
| Sex |  |  |  |  |
| Male | 107 (47.3) | 236 (54.9) | Ref. | Ref. |
| Woman | 119 (52.7) | 194 (45.1) | 1.35 (0.97,1.87) | 1.47 (1.04,2.08) |
| Income<sup>4</sup> |  |  |  |  |
| Under \$50,000 | 55 (24.3) | 132 (30.7) | Ref. | Ref. |
| \$50,000 to \$100,000 | 49 (21.7) | 98 (22.8) | 1.20 (0.76,1.91) | 1.17 (0.73,1.86) |
| \$100,000 to \$150,000 | 28 (12.4) | 39 (9.1) | 1.72 (0.98,3.07) | 1.4 (0.81,2.41) |
| Over \$150,000 | 22 (9.7) | 44 (10.2) | 1.20 (0.66,2.18) | 1.25 (0.7,2.28) |
| Race<sup>5</sup> |  |  |  |  |
| White | 104 (46.0) | 163 (38.0) | Ref. | Ref. |
| Hispanic | 53 (23.5) | 146 (34.0) | 0.57 (0.38,0.85) | 0.72 (0.46,1.12) |
| Asian | 7 (3.1) | 58 (13.5) | 0.19 (0.08,0.44) | 0.24 (0.1,0.55) |
| Black | 20 (8.8) | 18 (4.2) | 1.74 (0.88,3.44) | 2.54 (1.24,5.15) |
| More than 1 race | 26 (11.5) | 36 (8.4) | 1.13 (0.64,1.97) | 1.4 (0.78,2.51) |
| Native American or Alaskan Native | 6 (2.7) | 4 (0.9) | 2.34 (0.64,8.48) | 2.02 (0.54,7.53) |
| Native Hawaiian or Pacific Islander | 3 (1.3) | 1 (0.2) | 4.73 (0.48,42.82) | 4.64 (0.46,45.74) |

Logistic regression models adjusting for age, region, sex, income, and race predicted the likelihood an individual was vaccine hesitant. Missing values of income and race were multiply imputed using the Amelia II package.
<sup>1</sup>Case status is presented here for context but was not included in regression analyses as it could be considered an outcome of willingness to receive vaccination.
<sup>2</sup>Predominantly urban regions include San Francisco Bay Area, Greater Los Angeles Area, Greater Sacramento area, San Diego and the Southern border. We tabulate regions of residence for individuals who were hesitant or willing to receive vaccination in **Table S1**.
<sup>3</sup>Predominantly rural regions include Central Coast, Northern Sacramento valley, San Joaquin Valley, Northwestern California, and the Sierras region. We tabulate regions of residence for individuals who were hesitant or willing to receive vaccination in **Table S1**.
<sup>4</sup>For regression analyses, values were imputed for individuals who did share income data due to refusal (43 [19.0%] among hesitant and 66 [15.3%] among non-hesitant participants) or those who did not know their income (29 [12.8%] among hesitant and 51 [11.9%] among non-hesitant participants).
<sup>5</sup>For regression analyses, values were imputed for individuals who did not share race data (7 [3.1%] among hesitant and 3 [0.7%] among non-hesitant participants).

**Table 4:** Reasons for vaccine hesitancy among individuals not yet vaccinated.

| Stated reason | n (%) among 219 respondents reporting hesitancy to receive vaccination |
| --- | --- |
| Concerned about any vaccine side effects | 66 (30.0) |
| Concerned about long term vaccine side effects | 60 (27.4) |
| Concerned about COVID-19 vaccine safety | 60 (27.4) |
| Waiting to see more research on COVID-19 vaccines | 40 (18.3) |
| I have not yet thought about whether I want the COVID-19 vaccine | 24 (11.0) |
| Currently infected with SARS-CoV-2 | 23 (10.5) |
| Concerned about safety for vaccines generally | 22 (10.0) |
| Do not believe vaccination against COVID-19 is important | 20 (9.1) |
| Not at high risk for COVID-19 | 17 (7.8) |
| Currently pregnant | 9 (4.1) |
| Do not trust the government | 9 (4.1) |
| Negative reaction to prior vaccinations | 5 (2.3) |
| Lack of trust in the medical system | 5 (2.3) |
| Would only get vaccine if required by school/work | 5 (2.3) |
| Contraindicated medical condition | 5 (2.3) |
| Afraid of getting SARS-CoV-2 from the vaccine | 3 (1.4) |
| Depends on the vaccine product offered | 2 (0.9) |
| Object to vaccination due to religious reasons | 2 (0.9) |
| Afraid of needles | 1 (0.5) |

<sup>1</sup>Calculated out of N=219 because 7 individuals declined to answer.

## DISCUSSION

Among a sample of the general population of Californians, available mRNA-based COVID-19 vaccines demonstrated robust protection against documented SARS-CoV-2 infection under real-world conditions. While we identified partial protection before two weeks after receipt of the second dose, similar to other published estimates [7,9], the increase in VE from 68% following the first dose to 87% at ≥15 days after receipt of the second dose corresponds to a 59% incremental reduction in risk. We also found that mRNA-based COVID-19 vaccines elicit substantial protection against both symptomatic infections and infections for which participants reported healthcare-seeking, with 91% VE against each of these endpoints. No hospitalizations were observed among fully vaccinated cases within our study, consistent with findings of other published studies demonstrating strong protection against clinically-severe COVID-19 endpoints [12]. Our results closely resemble estimated efficacy of mRNA-based COVID-19 vaccines in trials that monitored for symptomatic COVID-19 endpoints [1,2]. Moreover in our study, the low frequency of post-vaccination infections, and our estimate of 68% VE against infections for which participants did not report symptoms, indicates vaccination may substantially reduce SARS-CoV-2 virus circulation within the community.

Our finding that 66% of unvaccinated participants in this early period of vaccine rollout were willing to receive COVID-19 vaccination align with national estimates of COVID-19 vaccine confidence [13]. We further identified rural-urban divides in vaccine enthusiasm, in addition to lower vaccine confidence among female and Black participants. Concerns over vaccine safety and side effects were reported by only a minority of all participants who expressed hesitancy about receiving COVID-19 vaccination (27-30%), but were the most commonly cited reasons for hesitancy. Recent studies have documented emerging differences in acceptance of COVID-19 vaccination associated with region of residence, educational background, employment status, and ideological factors [14–16]. Differing messaging and outreach strategies will thus be needed to address barriers to vaccine acceptance across communities, including people whose hesitancy to receive vaccination stems from mistrust or adverse experiences within US healthcare systems [17]. Prior studies have demonstrated that a provider’s recommendation is a key determinant of vaccine acceptance [18]. As healthcare providers in California and other settings have generally reported high (although not universal) enthusiasm around receiving COVID-19 vaccination [19,20], they may serve as important advocates to encourage vaccine uptake in their communities.

Limitations of our study should be considered. While observational studies face risks of bias, similarity of our estimates to those of other studies, stepwise increases in VE with time since receipt of each dose, and the absence of apparent protection immediately following first-dose receipt each support external validity of our findings [7,8,21]. Reliance on participants being available and willing to answer the phone is a limitation, although this applied to both cases and controls who received SARS-CoV-2 testing.

Nonetheless, our study may have under-enrolled participants experiencing very severe illness (e.g. who are hospitalized, have died, or are unable to participate in the phone interview due to sickness), who would be unable to answer the phone. As such, the findings should be interpreted as estimates of VE against a primarily mild to moderate spectrum of illness. We did not identify differential willingness to participate in the study among persons who tested positive and negative, provided contact was made. While misclassification of self-reported vaccination is possible, we did not find significant differences in VE estimates between analyses that did or did not restrict data to include participants who referenced a vaccine card. We did not re-contact cases to verify that cases who reported no symptoms remained asymptomatic over the course of their infection, or to confirm that cases who were not hospitalized or had not sought advice from healthcare providers at the time of their interview did not subsequently receive such care. Last, it is possible that certain participants were unaware of prior SARS-CoV-2 infections they may have experienced, particularly if these infections were mildly symptomatic or asymptomatic. Immunity resulting from such infections could lead to lower estimates of VE under our study design [6,22].

Our findings indicate that vaccine rollout is preventing COVID-19 in the general population of California and significantly reducing the risk of both asymptomatic and symptomatic SARS-CoV-2 virus infection. Vaccine hesitancy among historically marginalized and rural populations, which account for a substantial proportion of all COVID-19 cases in California to date [4], presents a barrier to reaching coverage levels needed for herd immunity.

## Data Availability

Requests for data should be communicated to the California Department of Public Health.

## FUNDING

The study was supported by the California Department of Public Health. JP, JO, and JFM were supported by a grant from the ELC program of the US CDC (program number 0187.0150). JAL and NPJ were supported by NIH/NIAID (grant R01-AI14812701).

## ACKNOWLEDGMENTS

Members of the California COVID-19 Case-Control Study Team include: Helia Samani, Sophia S. Li, Camilla M. Barbaduomo, Nikolina Walas, Christine Wan, Anna T. Fang, Timothy Ho, Vivian H. Tran, Erin Xavier, Mahsa H. Javadi, Diana J. Poindexter, Najla Dabbagh, Michelle M. Spinosa, Nozomi Birkett, Paulina M. Frost, Zheng N. Dong, Shrey Saretha, Adrian F. Cornejo, Jennifer L. DeGuzman, Miriam I. Bermejo, Hyemin Park, Amanda Lam

## CONFLICTS OF INTEREST

JAL discloses receipt of grants and honoraria from Pfizer, Inc. unrelated to this study.

## Supplementary information

**File S1. Survey Questionnaire**

### SECTION 0: REGISTER THAT THE CALL IS PLACED

1. Please select your first and last name
2. Please paste the case-control ID. Note: this is the “LinkLog” in column A of the spreadsheet
3. Please select the region for the case-control:
4. Please select the sex of the case-control:
5. Please select the age of the case-control you are interviewing:
6. Please write the date that the case-control test was administered (MM/DD/YYYY)
7. Please write the date that occurred 14 days prior to the date above (MM/DD/YYYY)

*Please select “Nobody answered the phone” if the call did not go through, nobody answered the phone, or the call went to voicemail after TWO attempts at each number provided. Otherwise, proceed with introductions:*

### SECTION 1: INTRODUCTION (∼2 min)

1. **Hello, my name is [**_______**] and I am calling on behalf of California Department of Public Health to ask some questions regarding [NAME]’s recent COVID-19 test on [INSERT DATE OF TEST]**.
2. *Make sure you’re on the phone with the correct person*. *If case is a child under 18y, make sure you are speaking to a parent/ guardian:*
  **2a. Am I speaking to [NAME]’s parent or guardian?** [If yes, proceed to **section 2**] [If no, proceed to 2b]
  **2b. Can you please pass the phone to [NAME]’s parent or guardian?** [If yes, proceed to **section 2**] [If no, end call] *If case is someone older than 18y:*
  **2c. Am I speaking to [NAME]?** [If yes, proceed to **section 2**] [If no, proceed to 2d]
  **2d. Can you please pass the phone to [NAME]?** [If yes, proceed to **section 2**] [If no-end call]

*NOTE on proxy respondents:*

*If an individual is hospitalized or otherwise too sick to answer questions on their own behalf, a caretaker may serve as a proxy respondent, but verbal consent must first be obtained from the primary case both to participate in the study and to have the proxy respondent answer on their behalf*.

*A proxy respondent who speaks English or Spanish may answer if the individual is unable to easily complete the interview in one of these two languages, provided they are able to speak English or Spanish with sufficient proficiency to provide verbal consent for both participation and for communicating via the proxy respondent*.

### SECTION 2: ASSENT (∼1 min)

*If you are speaking to a parent or correct person for the first time, add your name and affiliation before starting:*

**Hello, my name is [_____]and I am calling on behalf of California Department of Public Health**.

1. **Hi! We are interested in asking you some questions about [YOUR or INSERT CHILD’S NAME] recent COVID-19 test. We are hoping to interview you to try to better understand the spread of COVID-19. Do you have some time to chat?** INTERVIEWER: pause and wait for person to confirm that they are still on the line, check YES if they say they are willing to chat If they do not have time, select NO
2. **So before we start, I want to make sure you understand that everything I ask you is confidential, protected by California’s strict privacy laws, and is only being used to inform public health. Your answers will not be shared with any other federal, state, or local authorities, and you’re welcome to decline to answer any question. We anticipate this will take about 20 minutes. I know that sounds like a long time, but we really appreciate your time and your answers will help us answer some extremely important questions about COVID-19**. **Do you understand the information I have just shared with you?** INTERVIEWER: check “yes” if the respondent answers yes and if you deem the respondent to be competent to proceed with consent and interviewing; check “no” and thank the respondent for their time if the respondent says no or if you deem the respondent is not competent to proceed with consent and interviewing.] If it seems like the person needs a<u>proxy respondent</u> due to not speaking well enough English or being too sick, you may ask “**Is there anyone who can help you answer my questions?”**. If you get the proxy respondent on the phone, re-introduce yourself by starting at the top of Section 2 with “Hello, my name is…” and add at the end, **“Can you help answer questions on [insert name of case/control’s] behalf?”** NOTE that a proxy respondent must be over the age of 14. *Interviewers then seek consent from the participant, but the question asked will depend on the age of the desired case/control*. [If participant is answering on their own behalf AND they are older then 18] **Great, thank you! To confirm, are you willing to participate in this interview?** [If participant is a child older than 14, answering on their own behalf, first ask for consent from the parent for the child to answer the survey] **Great, thank you! I want to let you know that your child [INSERT CHILD’S NAME] may answer questions on their own behalf. Are you willing to allow [INSERT CHILD’S NAME] to participate in this interview? If not, you can answer questions on their behalf**. *Interviewer: if the child older than 14 joins the call, make sure to reintroduce yourself and explain the purpose of the survey*. [If participant is a child younger than 14, and adult is answering on their behalf] **Great, thank you! Are you willing to answer questions about [INSERT CHILD’S NAME]’s recent exposures as part of this interview?** [If a proxy respondent will answer on behalf of the study participant] **If you are able, I would suggest putting the phone on speakerphone during this interview, so [insert name/ relationship of proxy respondent] can help you**. **[INSERT NAME OF CASE-CONTROL], are you willing to participate in this interview?** **[INSERT NAME OF CASE-CONTROL], do you consent to allow [NAME OF PROXY RESPONDENT] to answer my questions during this interview. Please stay close by [NAME OF PROXY RESPONDENT] in case it is necessary to clarify any points that come up**. [If no or asks to be called back later, proceed to end of the survey] [If consent is provided and case/control is 7-18 years old, proceed to 3] *Interviewer: select the following options based off of the consent pattern:*
  - Participant provided consent on their own behalf
  - Parent provided consent for child <18 yrs
  - Participant provided consent for proxy respondent to answer on their own behalf
  - No consent was provided
3. **No problem. But before we hang up, do you mind quickly sharing why you are unable or unwilling to complete this call?** *Record the free response* [**<u>End call</u>**]
4. **[INSERT CHILD’S NAME] is welcome to stand by or join the call to help answer questions**. [If child joins the call, proceed to 3b, otherwise skip to **section 3**] **3b. Hi [INSERT CHILD’s NAME]. My name is [] and I work with the California Department of Public Health. I’m going to ask you some questions about activities in the past couple of weeks. Are you willing to answer these questions so that we can better understand the spread of COVID-19?** [Proceed to **section 3**]

### SECTION 3: LAST COVID TEST (∼3 min)

1. **Great, so to start, I want to ask whether you know your COVID-19 test result from [INSERT DATE OF TEST]?** *Record whether they know or don’t know their test result by selecting on of the options:*
  - Subject knows test result and is positive
  - Subject knows test result and is negative
  - Subject does NOT know test result and is positive
  - Subject does NOT know test result and is negative [If yes and they are positive, proceed to **section 4**] [If yes and they are negative, proceed to 3] [If no, and they are negative, proceed to 2] [If no, and they are positive, proceed to 4]
2. **Your COVID-19 test result from [INSERT DATE OF TEST] has come back negative**. *Record one of the following options:*
  - Yes
  - No
  - Don’t know
  - Refuse [Proceed to 3]
3. **Have you ever received a *positive* COVID-19 test result or been told by a health care provider that you are positive for COVID-19?** [If no, proceed to **section 4**] [If yes, end-call saying: **Thanks for letting me know. Those are all the questions I have for you. Thank you for your time and I hope you have a nice day**.
4. **Your COVID-19 test result from [INSERT DATE OF TEST] has come back positive. This means you do have coronavirus disease or COVID-19. In my role with CDPH, I cannot provide you with medical advice. If you need any medical information, please call your healthcare provider. One thing I want to be sure of today is that we have a plan for you to follow up with your healthcare provider, so that they can check on any symptoms you may have and assess your risks. Even if you feel okay now, it is important to have someone you can call if you start feeling sick. If you do not have a healthcare provider, you can go to an urgent care facility or the emergency room if you are not getting better or you feel like you are getting worse**. [Proceed to **section 4**] [If the person brings up clinical questions or concerns about their positive test] **Thank you for sharing that concern. In my role with CDPH, I am not able to give you medical advice. I do want to be sure that you get the help you need. If you believe you are having a medical emergency, you should call 911. Some warning signs that you should go to the emergency room for are: trouble breathing, bluish lips or face, pain or pressure in the chest that does not go away, new confusion or trouble waking or staying awake, but there are other symptoms too. Otherwise, you should call your healthcare provider**.

### SECTION 4: REASONS FOR TESTING (∼3 min)

1. **Next, I’m going to ask you some questions about your COVID-19 test. Can you describe to me why did you choose to get tested on [INSERT DATE OF TEST]?** *Interviewers will select check boxes from the respondent based off of their response, without prompting them from the following list, and will use a write-in option for any additional reasons for seeking testing. After choosing the best answer from the list, confirm your choice the case/control (ex. “So you got tested for pre or post-travel screening?”)*
  - I had contact with someone who tested positive
  - I had contact with someone who had symptoms, but I do not know if they were confirmed to be positive
  - I was told by a public health worker to get tested because I was exposed to a case
  - I was concerned about symptoms I experienced
  - Someone in my household had contact with someone who was positive
  - A person in my household had contact with someone who had symptoms or suspected they had COVID, but we do not know if they are confirmed to be positive.
  - Routine screening for my job
  - Pre or post-travel screening
  - Test required for a medical procedure
  - I just wanted to see if I was infected
  - Don’t know
  - Refuse
  - Other [*interviewer writes in response*]
2. **At the time you were tested on [DATE OF TEST] were you experiencing any COVID-19 symptoms?** *Record Yes/No/Not Sure/ Refuse* [If yes, ask <u>question 4</u>] [If no, proceed to <u>question 5</u>]
3. **Can you please list the symptoms you were experiencing on or 14 days prior to your test on [DATE OF TEST]** *Interviewers will select the symptoms the individuals indicated that they were experiencing. When the respondent is done listing symptoms, the interviewer may prompt, “Are you sure those were all the symptoms you experienced?” and proceed to confirm absence of the 6 most common symptoms (as applicable), in a conversational manner: “No fever, no chills, no muscle pain, no loss of appetite, no shortness of breath, no cough?”* *Select from the following list of symptoms:*
  - Blocked nose
  - Chills
  - Cough
  - Chest pain
  - Diarrhea
  - Muscle pain
  - Fever
  - Headache
  - Hoarseness
  - Loss of appetite
  - Loss of taste
  - Loss of smell
  - Myalgia (muscle pain)
  - Nausea
  - Runny nose
  - Shortness of breath
  - Sneezing
  - Sore throat
  - Stomach pain
  - Sinus pain
  - Sweating
  - Swollen glands
  - Tickle in throat
  - Watery eyes
  - Don’t know
  - Refuse
  - Other
4. **I am now going to read a list of places you may have sought treatment or advice prior to your test on [DATE OF TEST]. After I read the following options, please answer “Yes” or “No”**. *Record Yes/No/ Not Sure/Refuse e for each of the options below*
  - **Did you seek care at an in-person appointment with your usual physician or healthcare provider**
  - **Did you seek care at a telehealth visit or phone appointment with your usual physician or healthcare provider**
  - **Did you seek care at an in-person visit to an urgent care clinic**
  - **Did you seek care at an in-person visit to a healthcare provider at a retail pharmacy**
  - **Did you visit the emergency room?**
  - **Were you admitted to the hospital?**
  - **And just to follow-up, where there any other forms of healthcare from which you sought treatment advice at the time you had your test on [Insert date of test](specify):_______**
5. **In the 14 days prior to your test (between ADD DATE to ADD DATE) do you know whether you had known or suspected contact with one or more people who may have tested positive for COVID-19?** *Select one of the following options*
  - Yes-contact with one person who was confirmed positive
  - Yes-contact with more than one person who was confirmed positive
  - Yes-contact with one person who I suspected was positive
  - Yes-contact with more than one person who I suspected was positive
  - No known or suspected contact with a positive case
  - Not sure
  - Refuse [If case indicated they had KNOWN or SUSPECTED Contact, proceed to **section 5**, **<u>part A</u>**], [If the case did not have known or suspected contact, proceed to **section 6**]

### SECTION 5: CONTACT WITH KNOWN OR SUSPECTED CASE (∼8 min)

[If case indicated they had KNOWN or SUSPECTED Contact, proceed to A]

[If case indicated they did NOT have known or suspected contact, proceed to **section 6**]

**A. I’m going to now ask you some questions about the type of contact you had with the person (people) who may have had COVID-19. We are trying to understand sources of exposure and are hopeful that you are willing to answer the questions honestly, knowing that we aren’t looking or expecting any sort of answer**.

1. **Was the known/ suspected contact someone who lives in your household?** *if plural (contact with >1 person*): **Were any of the known/ suspected contact people who lives in your household** *Record Yes, No, Don’t know, Refuse*
2. **Did the known/ suspected contact occur indoors, outdoors, or both indoors and outdoors?** *if plural (contact with >1 person):* **Did the known/suspected contacts occur indoors, outdoors, or both indoors and outdoors?** *Record Indoors, Outdoors, Both indoors and outdoors, Unknown, or Refuse*
3. **In the 14 days prior to your test (between [ADD 14 DAYS – TEST DATE HERE] to [ADD TEST DATE]), what are the locations where you may have had contact with this person?** *if plural:* **In the 14 days prior to your test (between [ADD 14 DAYS – TEST DATE HERE] to [ADD TEST DATE]), what are the locations where you may have had contact with these people)** *Record the free response answer*
4. **I am now going to ask you about different precautions you may or may not have been able to take when you came into contact with the known or suspected positive case. Please answer “Yes, No or Not Sure” after each question:** *Record Y/ N/ Not sure for each of the options below:*
  - **Did you come within 6 feet of this person, indoors?** *If plural:* **Did you come within 6 feet of any of these people, indoors?**
  - **Did you come within 6 feet of this person, outdoors?** *If plural:* **Did you come within 6 feet of any of these people, outdoors?**
  - **Did you have physical contact with this person, (ie. handshake, hug)?** *If plural*: **Did you have physical contact with any of these people (ie. handshake, hug)**
5. **Did you wear a mask the entire time, most of the time, some of the time, or none of the time that you interacted with this person?** *If plural*: **Did you wear a mask the entire time, most of the time, some of the time, or none of the time that you interacted with these people?** *Record which of the statements they agree with from below:*
  - I wore a mask the entire time I interacted with this (these) person(s)
  - I wore a mask most of the time I interacted with this (these) person(s)
  - I wore a mask some of the time I interacted with this (these) person(s)
  - I did not wear a mask during this (these) interaction(s)
  - Not sure
  - Refuse
6. **Did the person you had known or suspected contact with wear a mask the entire time, most of the time, some of the time, or none of the time when you interacted with them?** *If plural:* **Did the people you had known or suspected contact with wear a mask all, most, some, or none of the time that you interacted with them** *Record which of the statements they agree with from below:*
  - They wore a mask the entire time we interacted
  - They wore a mask most of the time we interacted
  - They wore a mask some of the time we interacted
  - They did not wear a mask during this interaction
  - Not sure
  - Refuse
7. **Did you spend more than 3 consecutive hours with this person in the 14 days prior to your test (between Date to Date)**. *If plural*: **Did you spend more than three consecutive hours with these people in the 14 days prior to your test (between Date to Date)** *Record Yes/ No/ Don’t know/ Refuse* [proceed to **section 6**]

### SECTION 6: EXPOSURE WITH CONTACT KNOWN OR SUSPECTED CASE (∼10 min)

**Next, I want to learn about potential sources of exposure to COVID-19 in the 14 days before your last test: from [ADD 14 DAYS – TEST DATE HERE] to [ADD TEST DATE]. It may help you to pull up a calendar to remember what you were up to over the last two weeks. This chunk usually takes the longest, so thank you in advance for your time**

*Only read the following if they did <u>not</u> have known or suspected contact:*

**[We are trying to understand sources of exposure and are hopeful that you are willing to answer the questions honestly, knowing that we aren’t looking or expecting any sort of answer.]**

1. **I am now going to ask you about a series of locations which you may have visited. After I announce each location, please tell me “Yes, No, or Not sure” to indicate whether you visited that location between [ADD 14 DAYS – TEST DATE HERE] to [ADD TEST DATE]**.
  - **First, did you attend a health appointment or health facility (other than where you got tested for COVID-19)**
  - **Did you go grocery shopping?**
  - **Now I am going to ask you about the times you went to restaurants. Did you go to any restaurants to pick up take-out or to eat at the restaurant?** *Record one of the following options: a) Dine-in (eat at restaurant) only, b) Take-out only, c) Both dining-in and take-out, d) Neither dine-in or take-out, e) Not sure, f) Refuse* *If yes and take-out:*
  - **How many times did you get take-out?**
  - **Did you ever have to go inside the restaurant to either place or pick up your take-out order?** *Record one of the following options: a) Yes, I went inside the restaurant either to place or pick-up my order, b) No I did not go inside the restaurant either to place or pick-up my order, d) No I did not go inside the restaurant either to place or pick-up my order, but someone I went to the restaurant with had to go inside to place or pick-up the order, e) not sure, f) refuse* *If yes and dine-in:*
  - **How many times did you eat at an <u>indoor</u> restaurant?**
  - **How many times did you eat at an <u>outdoor</u> restaurant?** [Skip the following question chunk about bars if respondent is under 21]
  - **Did you attend any bars, breweries, or wine bars?** *If yes, ask:* **Did you attend a bar, brewery or wine bar?** *Select all For each of the places they indicated that they visited:*
  - **How many times did you attend a [bar/brewery/wine bar]?**
  - **When you went to a (those) [bar(s)/brewery(ies)/wine bar(s)], did you spend most of your time indoors, outdoors, or both indoors and outdoors?**
  - **Did you ever visit a coffee shop?** *If yes, ask:*
  - **When you (typically) visited the coffee shop(s), did you have to go inside to place your order?** *Record one of the following options: a) I went inside to place the order, b) I typically placed the order outside or remotely (via. App, web portal, phone order), c) Don’t know, d) refuse*
  - **When you visited a coffee shop, did you (typically) consume your beverage inside the shop, outside the shop, or did you just pick-up the beverage for take-away**. *Record one of the following options a) consumed inside the shop, b) consumed outside the shop (ex. restaurant set up outdoor tables/ chairs and I drank/ate at those tables), c) Got beverage for take-away, d) Don’t know, e) refuse*
  - **Did you go retail shopping?** *If yes, ask:* **And did you go indoor or outdoor retail shopping?**
  - **Did you exercise at gym?** *If yes, ask:* **And was this an indoor or an outdoor gym?**
  - **Did you participate in a group recreational sport (tennis, soccer, basketball, swimming)**
  - **Did you ever leave your house to go for a walk, run, hike or ride a bike outside?** *If yes ask:* **Did you hike, run, walk, or bike with anyone outside your household?** *Select one of the following options a) No, I always hiked, ran, walked, or biked by myself, b) No, but I sometimes/ always ran, walked, or biked with other people who live in my household, c) Yes I hiked ran, walked, or biked with someone who doesn’t live in my household, d) Don’t know, e) Refuse*
  - **Did you ride public transit?**
  - **Did you use a ride share (eg. Taxi, Uber, Lyft, or carpool with individuals who are not members of your household) ?**
  - **Did you fly on a plane?**
  - **Did you attend a parade, rally, march, or protest?**
  - **Did you receive services at a salon or barber?**
  - **Did you attend an indoor movie theater?**
  - **Did you attend a worship service?** *If yes ask:* **And was this an indoor or an outdoor worship service?**
  - **Did you visit or stay at a school, daycare or preschool?** *If yes, ask:* **Was the school or daycare public or private?**
  - **Did you visit a jail, prison, or correctional facility?** *If a participant answers yes to any of the questions in 1, follow-up with:* **How many times did you attend [INSERT LOCATION] between [ADD 14 DAYS – TEST DATE HERE] to [ADD TEST DATE]**. **I am now going to ask you (a couple more) some questions about face mask usage between date to date**.
2. **Between [ADD 14 DAYS – TEST DATE HERE], at all of the <u>indoor</u> places we discussed earlier, did you wear a face mask all, most, some, or none of the time?**
  - I wore a face mask all of the time
  - I wore a face mask most of the time
  - I wore a face mask some of the time
  - I never wore a face mask in indoor places
  - I did not go inside any indoor places other than my home
  - I was not in contact with anyone
3. **Between [ADD 14 DAYS – TEST DATE HERE], at all of the <u>indoor</u> places we discussed earlier, did people you came within 6 feet of wear a face mask all, most, some, or none of the time?**
  - They wore a face mask all of the time
  - They wore a face mask most of the time
  - They wore a face mask some of the time
  - They never wore a face mask in indoor places
  - I did not go inside any indoor places other than my home
  - I was not in contact with any people outside my household in indoor places
4. **Between [ADD 14 DAYS – TEST DATE HERE], at all of the <u>outdoor</u> places we discussed earlier, did you wear a face mask all, most, some, or none of the time?**
  - I wore a face mask all of the time
  - I wore a face mask most of the time
  - I wore a face mask some of the time
  - I never wore a face mask in indoor places
  - I did not go inside any outdoor places other than my home
5. **Between [ADD 14 DAYS – TEST DATE HERE], at all of the <u>outdoor</u> places we discussed earlier, did people you came within 6 feet of wear a face mask all, most, some, or none of the time?**
  - They wore a face mask all of the time
  - They wore a face mask most of the time
  - They wore a face mask some of the time
  - They never wore a face mask in indoor places
  - I did not go inside any outdoor places other than my home
  - I was not in contact with any people outside my household in outdoor places
6. **I am now going to ask you some questions about social gatherings. These include any informal gatherings with friends or family who are NOT members of your household). Did you attend any social gatherings between (14 days prior to test result to test result date)?** *Interviewer: note that our definition of social gatherings is mixing with people who don’t otherwise live in your household. If someone had a longer-term family* together *(ie. traveled to visit relatives, but stayed for multiple days, count this as ONE event)*. *If yes, ask:* **When you attended social gatherings, were they indoors, outdoors, or both indoors and outdoors?** *An <u>outdoor only</u> gathering means the person spent the majority of their time outside An <u>indoor only</u> gathering means the person spent the majority of their time* *A gathering that was “<u>both indoors and outdoors</u>” means the participant was both inside and outside during the social gathering (ex. Sandy had some friends over for dinner and they ate outside on the patio, and then watched a movie in their living room together)* *If they indicate they attended indoor social gatherings***: How many indoor social gatherings did you attend between (14 days prior to test result to test result date)? About how many people attended these gatherings? Did you eat or drink during any of these (or this) gatherings? When you attended this (these) indoor gathering(s), did you wear a face mask all, most, some or none of the time?** *Interviewer: note that this question about mask usage is distinct from the question earlier*. *If they indicate they attended outdoor social gatherings:* **How many outdoor social gatherings did you attend between (14 days prior to test result to test result date)? About how many people attended these gatherings? Did you eat or drink during any of these (or this) gatherings? When you attended this (these) outdoor gathering(s), did you wear a face mask all, most, some or none of the time?** *If they indicate they attended social gatherings that were both inside and outside:* **How many social gatherings did you attend between (14 days prior to test result to test result date) that were both indoor and outdoor? About how many people attended these gatherings? Did you eat or drink during any of these (or this) gatherings? When you attended this (these) outdoor gathering(s), did you wear a face mask all, most, some or none of the time?**
7. **Did you attend any other kind of event where there are <u>5 or more</u> people who are not in your household in attendance?** *Interviewer: If necessary, prompt with options like a sporting event, concert, festival, etc*. **Specify the event:**

[Proceed to **section 7**]

### SECTION 7: OCCUPATION (∼1 min)

1. **I am now going to ask you some questions about your occupation. Between [ADD 14 DAYS – TEST DATE HERE] to [ADD TEST DATE] did you attend work, school, or volunteering commitments exclusively at home, both at home and in “in-person”, or exclusively “in-person”**.
  - I work, study, and/or volunteer at home
  - I attend work, school, and/or volunteering “in-person”
  - I attend work, school, and/or volunteering both “in-person” and at home
  - I am not currently working, in school, or in a volunteer position. [If respondent is a student, skip question and just record “student”]
2. **Can you tell me what your job is?** (*Record open ended response]* [If they attend work, school, or volunteering commitments in person or both at home & in person, proceed to question 3, otherwise proceed to **Section 8**]
  3a. **Do you come into close contact (within 6 feet) of more than 10 people per day at work/school/volunteering?** *Record: Yes or No*
  3b. **Do you primarily attend work/school/volunteering indoors, outdoor, or both indoors and outdoors?** *Record: indoors, outdoors, or both*

[Proceed to **section 8**]

### SECTION 8: VACCINATION (∼2 min)

**I am now going to ask you some questions about the COVID-19 vaccine**.

1. **Do you have any conditions that might place you higher risk for COVID-19?** *Interviewers may prompt with examples such diabetes, high blood pressure, overweight, being immunocompromised if requested. Select options from list below*
  - Lung conditions: COPD, lung cancer, cystic fibrosis, moderate to severe asthma, pulmonary fibrosis
  - Heart disease
  - High blood pressure
  - Obesity
  - Overweight
  - Diabetes
  - Weakened immune system: organ transplant, cancer treatment, bone marrow transplant, HIV/AIDS, sickle cell anemia, thalassemia
  - Chronic kidney disease
  - Chronic liver disease
  - Pregnant (first, second, or third trimester)
2. **Have you received any doses of a COVID-19 vaccine?** *Record: Yes, No* [If they have not received any doses, skip to 2b, otherwise ask question 3]
  2b. **Do you plan to receive any doses of the COVID-19 vaccine?** *Record: Yes, No, not sure, refuse* [If they are not planning to receive any doses or are not sure yet, ask 2c, otherwise, ask skip to **section 9**]
  2c. **Can you describe to me why you are not planning to receive the COVID-19 vaccine?** *Record reason in check box*
3. **How many doses of the COVID-19 vaccine have you received?** *Record: 1, 2*
4. **Do you have a vaccine card on hand from when you got the COVID-19 vaccine?** *If yes, ask them to get their vaccine card. If no, ask them to do their best remembering and try pulling up a calendar to help them remember*.
5. **What dates did you receive your dose(s)?** *Record the date of each vaccine*
6. **Do you know what product COVID-19 vaccine you received?** *Record the product of each dose*
7. **Do you have access to a COVID -19 vaccination clinic at your work or school?** *Record: yes/ no/ not sure/ refuse*
8. **Where did you get your COVID-19 vaccine?** *Record: mass vaccination site, hospital, nursing home, at my work, at my school, at a retail pharmacy, at a retail shop (eg. Walmart)*
9. **At the time you received the vaccine was it required to attend work or school?** *Record: yes/ no/ not sure/ refuse*

[Proceed to **section 9]**

### SECTION 9: DEMOGRAPHICS (∼5 min)

**I just have a few more questions. Again, anything you share with me is confidential and protected by California’s strict privacy laws. The information we collect about you will assist the health department in their COVID-19 response**.

1. **First, I’m going to ask you some general questions about COVID-19. From the beginning of the pandemic to the time you were tested on [DATE OF TEST], how worried did you feel about getting COVID-19? Would you say you felt:**
  - **Very worried**
  - **Somewhat worried**
  - **Neutral**
  - **Not worried at all**
2. **Since the beginning of the pandemic, there have been a lot of recommendations on behaviors that can reduce the risk of COVID-19 including avoiding large crowds, travel, and maintaining 6 feet of distance in public places. Would you say that you strongly agree, agree, are neutral, disagree, or strongly disagree that these measures reduce the risk of COVID-19?** *Record strongly agree, agree, neutral, disagree, strongly disagree*
3. Another recommendation to reduce the spread of COVID-19 is wearing face masks. Would you say that you strongly agree, agree, are neutral, disagree, or strongly disagree that face masks reduce the risk of COVID-19? *Record strongly agree, agree, neutral, disagree, strongly disagree*

**Last, I want to capture some information about demographics**.

1. **So do you mind sharing how old you are?** *Record free response*
2. **Next, please let me know which of the following race/ethnicities best describe yourself. You may select all that apply:**
  - **White**
  - **Black**
  - **Hispanic**
  - **Asian**
  - **Native American or Alaska Native**
  - **Native Hawaiian or other Pacific Islander**
3. **What is your sex/ gender?** *Record Man, Woman, Non=-binary, Prefer to self-describe, Refuse, Don’t know*
4. **What is your zip code of your home address?** *Record address using encryption tool*
5. **What is your home address?** *Record address using encryption tool after verifying it is an address using google maps*
6. **Which of the following best describes your living arrangement:**
  - **Private home**
  - **Apartment, or condominium**
  - **Skilled nursing facility**
  - **College or university student housing**
  - **Military quarters**
  - **Emergency or transitional shelter**
  - **Other (please describe)**
7. **How many people live in your household?**
8. **How many bedrooms do you have in your household?**
9. **Do you have any children under 18 at your home?**
10. **Are any of your children under 18 attending in-person instruction, school, or daycare?**
11. **Does anyone visit your home on a regular basis like a cleaning service or babysitter?** If you are talking to a child aged 14-17, at this point you can end the interview with the child and ask to speak with their parent/guardian. When you get back on the phone with the parent or guardian, you can say something like **[“Hi again, thank you so much for letting me speak with your child, it was extremely helpful. We are wrapping up the survey with some demographic questions and my last question that I didn’t want your child to have to answer was whether you are willing to share your total household income?”]**
12. **What is your total household income? Answer on behalf of everyone you share finances with**. [If you are speaking with POSITIVE case, proceed to 13] [If you are speaking with NEGATIVE control, proceed to 14]
13. **Thank you for participating in this survey. You may be contacted by another staff member at the health department to check in on you. They will ask you questions about your health and well-being to make sure you’re ok**.
14. **Thank you for participating in our survey. We appreciate your time**.

**Table S1:**
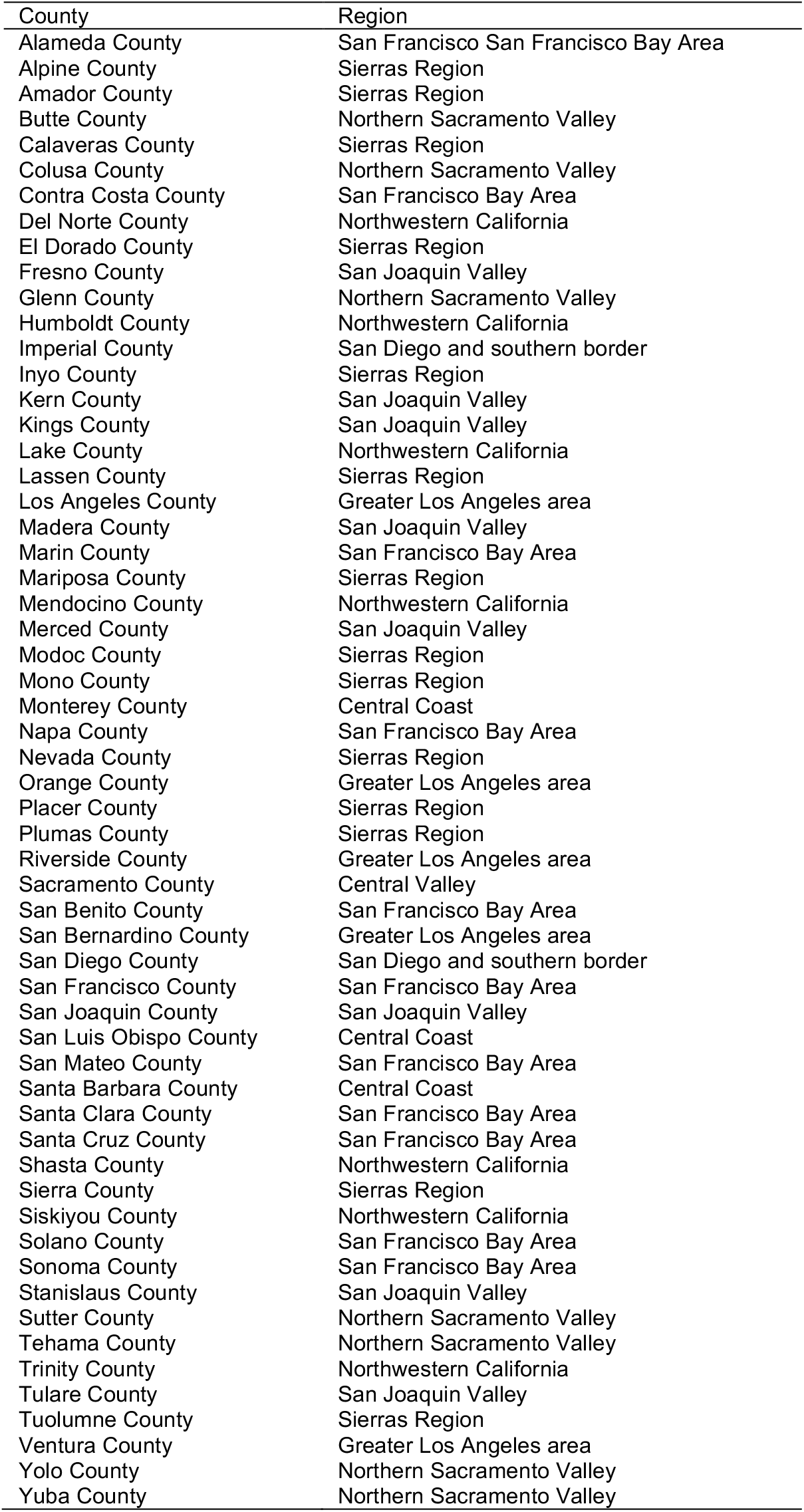
Counties by geographic region.

**Table S2:** Characteristics of total population eligible for inclusion.

|  |  | Eligible |  | Attempted |  | Final analytic sample |  |
| --- | --- | --- | --- | --- | --- | --- | --- |
|  |  | Case | Control | Case | Control | Case | Control |
|  |  | *n* (%) | *n* (%) | *n* (%) | *n* (%) | *n* (%) | *n* (%) |
|  |  | *N* = 108,606 | *N* = 4,718,559 | *N* = 3,847 | *N* = 5,253 | *N* = 525 | *N* = 498 |
| Age | <18 | 16,535 (15.2) | 650,959 (13.8) | 438 (11.4) | 540 (10.3) | -- | -- |
|  | 18-29 | 27584 (25.4) | 1179240 (25.0) | 1038 (27.0) | 1724 (32.8) | 200 (38.1) | 195 (39.2) |
|  | 30-49 | 34640 (31.9) | 1494701 (31.7) | 1219 (31.7) | 1584 (30.2) | 188 (35.8) | 175 (35.1) |
|  | 50-64 | 19918 (18.3) | 887593 (18.8) | 732 (19.0) | 956 (18.2) | 100 (19.0) | 92 (18.5) |
|  | 65+ | 9929 (9.1) | 506066 (10.7) | 419 (10.9) | 447 (8.5) | 37 (7.0) | 36 (7.2) |
| Region | *Predominantly urban regions* |  |  |  |  |  |  |
|  | San Francisco Bay Area | 25354 (23.3) | 1728944 (36.6) | 432 (11.2) | 694 (13.2) | 66 (12.6) | 63 (12.7) |
|  | Greater Los Angeles Area | 35706 (32.9) | 1728944 (31.5) | 537 (14.0) | 519 (9.9) | 48 (9.1) | 43 (8.6) |
|  | Greater Sacramento Area | 6412 (5.9) | 171474 (3.6) | 316 (8.2) | 576 (11.0) | 58 (11.0) | 57 (11.4) |
|  | San Diego and southern Border | 11190 (10.3) | 430201 (9.1) | 487 (12.7) | 545 (10.4) | 54 (10.3) | 56 (11.2) |
|  | *Predominantly rural regions* |  |  |  |  |  |  |
|  | Central Coast | 4352 (4.0) | 159802 (3.4) | 433 (11.3) | 778 (14.8) | 74 (14.1) | 66 (13.3) |
|  | Northern Sacramento Valley | 2334 (2.1) | 107760 (2.3) | 401 (10.4) | 482 (9.2) | 60 (11.4) | 56 (11.2) |
|  | San Joaquin Valley | 18398 (16.9) | 478466 (10.4) | 453 (11.8) | 596 (11.3) | 54 (10.3) | 52 (10.4) |
|  | Northwestern California | 1785 (1.6) | 59791 (1.3) | 345 (9.0) | 449 (8.5) | 55 (10.5) | 53 (10.6) |
|  | Sierras Region | 3075 (2.8) | 96366 (2.1) | 443 (11.5) | 614 (11.7) | 56 (10.7) | 52 (10.4) |
| Sex | Male | 53185 (48.9) | 2125671 (45.0) | 1937 (50.3) | 2705 (51.5) | 264 (50.3) | 255 (51.2) |
|  | Female | 55421 (51.1) | 2592888 (55.0) | 1910 (49.6) | 2548 (48.5) | 261 (49.7) | 243 (48.8) |

**Table S3:** Demographic attributes of vaccinated and unvaccinated cases and controls.

| Characteristics | Controls |  | Cases |  |
| --- | --- | --- | --- | --- |
|  | Unvaccinated, *n* (%)<br>*N*=313 | Vaccinated, *n* (%)<br>*N*=185 | Unvaccinated, *n* (%)<br>*N*=454 | Vaccinated, *n* (%)<br>*N*=71 |
| Age |  |  |  |  |
| 18-29 | 142 (45.4) | 53 (28.6) | 177 (39.0) | 23 (32.4) |
| 30-49 | 101 (32.3) | 74 (40.0) | 171 (37.7) | 17 (23.9) |
| 50-64 | 53 (16.9) | 39 (21.1) | 82 (18.1) | 18 (25.4) |
| 65+ | 17 (5.4) | 19 (10.3) | 24 (5.3) | 13 (18.3) |
| Region |  |  |  |  |
| *Predominantly urban regions* |  |  |  |  |
| San Francisco Bay Area | 35 (11.2) | 28 (15.1) | 54 (11.9) | 12 (16.9) |
| Greater Los Angeles Area | 34 (10.9) | 9 (4.9) | 40 (8.8) | 8 (11.3) |
| Greater Sacramento Area | 33 (10.5) | 24 (13.0) | 56 (12.3) | 2 (2.8) |
| San Diego and southern border | 37 (11.8) | 19 (10.3) | 49 (10.8) | 5 (7.0) |
| *Predominantly rural regions* |  |  |  |  |
| Central Coast | 41 (13.1) | 25 (13.5) | 63 (13.9) | 11 (15.5) |
| Northern Sacramento Valley | 35 (11.2) | 21 (11.4) | 52 (11.5) | 8 (11.3) |
| San Joaquin Valley | 36 (11.5) | 16 (8.6) | 43 (9.5) | 11 (15.5) |
| Northwestern California | 34 (10.9) | 19 (10.3) | 46 (10.1) | 9 (12.7) |
| Sierras Region | 28 (8.9) | 24 (13.0) | 51 (11.2) | 5 (7.0) |
| Sex |  |  |  |  |
| Male | 159 (50.8) | 96 (51.9) | 227 (50.0) | 37 (52.1) |
| Female | 154 (49.2) | 89 (48.1) | 227 (50.0) | 34 (47.9) |
| Household income |  |  |  |  |
| Under \$50,000 | 79 (25.2) | 40 (21.6) | 138 (30.4) | 15 (21.1) |
| \$50,000 to \$100,000 | 71 (22.7) | 36 (19.5) | 99 (21.8) | 14 (19.7) |
| \$100,000 to \$150,000 | 45 (14.4) | 31 (16.8) | 34 (7.5) | 11 (15.5) |
| Over \$150,000 | 31 (9.9) | 40 (21.6) | 50 (11.0) | 14 (19.7) |
| Refuse | 49 (15.7) | 19 (10.3) | 79 (17.4) | 7 (9.9) |
| Not sure | 38 (12.1) | 19 (10.3) | 54 (11.9) | 10 (14.1) |
| Race/Ethnicity |  |  |  |  |
| White | 128 (40.9) | 99 (53.5) | 178 (39.3) | 39 (54.9) |
| Hispanic | 86 (27.5) | 40 (21.6) | 143 (31.6) | 17 (23.9) |
| Asian | 33 (10.5) | 24 (13.0) | 50 (11.0) | 8 (11.3) |
| Black | 11 (3.5) | 6 (3.2) | 29 (6.4) | 1 (1.4) |
| More than 1 race | 43 (13.7) | 10 (5.4) | 32 (7.1) | 4 (5.6) |
| Native American | 4 (1.3) | 1 (0.5) | 10 (2.2) | 1 (1.4) |
| Native Hawaiian | 3 (1.0) | 3 (1.6) | 3 (0.7) | 1 (1.4) |
| Refuse | 5 (1.6) | 2 (1.1) | 8 (1.8) | 0 (0.0) |

**Table S4:** Frequency of each reported symptom and level of care sought by case-control status.

|  | Controls |  |  | Cases |  |  |
| --- | --- | --- | --- | --- | --- | --- |
|  | Unvaccinated,<br>n (%)<br>N=313 | Incompletely Vaccinated<sup>1</sup>,<br>n (%)<br>N=99 | Fully Vaccinated,<br>n (%)<br>N=86 | Unvaccinated,<br>n (%)<br>N=454 | Incompletely Vaccinated,<br>n (%)<br>N=51 | Fully Vaccinated,<br>n (%)<br>N=20 |
| **Symptoms** |  |  |  |  |  |  |
| Fever | 12 (3.8) | 4 (4.0) | 0 (0.0) | 120 (26.4) | 9 (17.6) | 2 (10.0) |
| Cough | 14 (4.5) | 4 (4.0) | 2 (2.3) | 134 (29.5) | 15 (29.4) | 2 (10.0) |
| Headache | 12 (3.8) | 4 (4.0) | 0 (0.0) | 141 (31.1) | 12 (23.5) | 1 (5.0) |
| Loss of taste | 2 (0.6) | 1 (1.0) | 0 (0.0) | 69 (15.2) | 4 (7.8) | 1 (5.0) |
| Loss of smell | 1 (0.3) | 1 (1.0) | 0 (0.0) | 66 (14.5) | 4 (7.8) | 1 (5.0) |
| Chills | 6 (1.9) | 1 (1.0) | 0 (0.0) | 75 (16.5) | 6 (11.8) | 0 (0.0) |
| Muscle Pain | 3 (1.0) | 2 (2.0) | 0 (0.0) | 91 (20.0) | 10 (19.6) | 1 (5.0) |
| Fatigue | 9 (2.9) | 1 (1.0) | 0 (0.0) | 81 (17.8) | 8 (15.7) | 2 (10.0) |
| Shortness of breath | 3 (1.0) | 2 (2.0) | 1 (1.2) | 41 (9.0) | 5 (9.8) | 0 (0.0) |
| Sore throat | 14 (4.5) | 4 (4.0) | 0 (0.0) | 69 (15.2) | 6 (11.8) | 2 (10.0) |
| Blocked nose | 8 (2.6) | 2 (2.0) | 0 (0.0) | 42 (9.3) | 7 (13.7) | 0 (0.0) |
| Runny nose | 10 (3.2) | 5 (5.1) | 1 (1.2) | 57 (12.6) | 6 (11.8) | 2 (10.0) |
| Chest pain | 3 (1.0) | 0 (0.0) | 1 (1.2) | 19 (4.2) | 1 (2.0) | 0 (0.0) |
| Watery eyes | 0 (0.0) | 0 (0.0) | 0 (0.0) | 4 (0.9) | 0 (0.0) | 0 (0.0) |
| Nausea | 3 (1.0) | 1 (1.0) | 0 (0.0) | 22 (4.8) | 0 (0.0) | 0 (0.0) |
| Sweating | 1 (0.3) | 0 (0.0) | 0 (0.0) | 9 (2.0) | 0 (0.0) | 1 (5.0) |
| Loss of appetite | 1 (0.3) | 1 (1.0) | 0 (0.0) | 25 (5.5) | 3 (5.9) | 0 (0.0) |
| Throat tickle | 1 (0.3) | 0 (0.0) | 0 (0.0) | 8 (1.8) | 1 (2.0) | 0 (0.0) |
| Any symptoms | 52 (16.6) | 13 (13.1) | 3 (3.5) | 354 (78.0) | 34 (66.7) | 9 (45.0) |
| **Level of care** |  |  |  |  |  |  |
| Hospital | 5 (1.6) | 3 (3.0) | 1 (1.2) | 15 (3.3) | 3 (5.9) | 0 (0.0) |
| Emergency room | 9 (2.9) | 3 (3.0) | 4 (4.7) | 37 (8.1) | 5 (9.8) | 1 (5.0) |
| Physician | 14 (4.5) | 2 (2.0) | 3 (3.5) | 16 (3.5) | 1 (2.0) | 0 (0.0) |
| Telehealth | 17 (5.4) | 4 (4.0) | 1 (1.2) | 67 (14.8) | 10 (19.6) | 1 (5.0) |
| Urgent care | 3 (1.0) | 2 (2.0) | 2 (2.3) | 23 (5.1) | 2 (3.9) | 0 (0.0) |
| Pharmacy | 6 (1.9) | 1 (1.0) | 0 (0.0) | 14 (3.1) | 0 (0.0) | 1 (5.0) |
| Any care<sup>2</sup> | 41 (13.1) | 11 (11.1) | 8 (9.3) | 132 (29.1) | 15 (29.4) | 3 (15.0) |

<sup>1</sup>An individual was considered incompletely vaccinated if they had received one or more doses of a mRNA COVID-19 vaccine product, but got tested <14 days after their second dose.
<sup>2</sup>Numbers for any care sought will not sum to the column totals owing to individuals who sought multiple forms of care

**Table S5:**
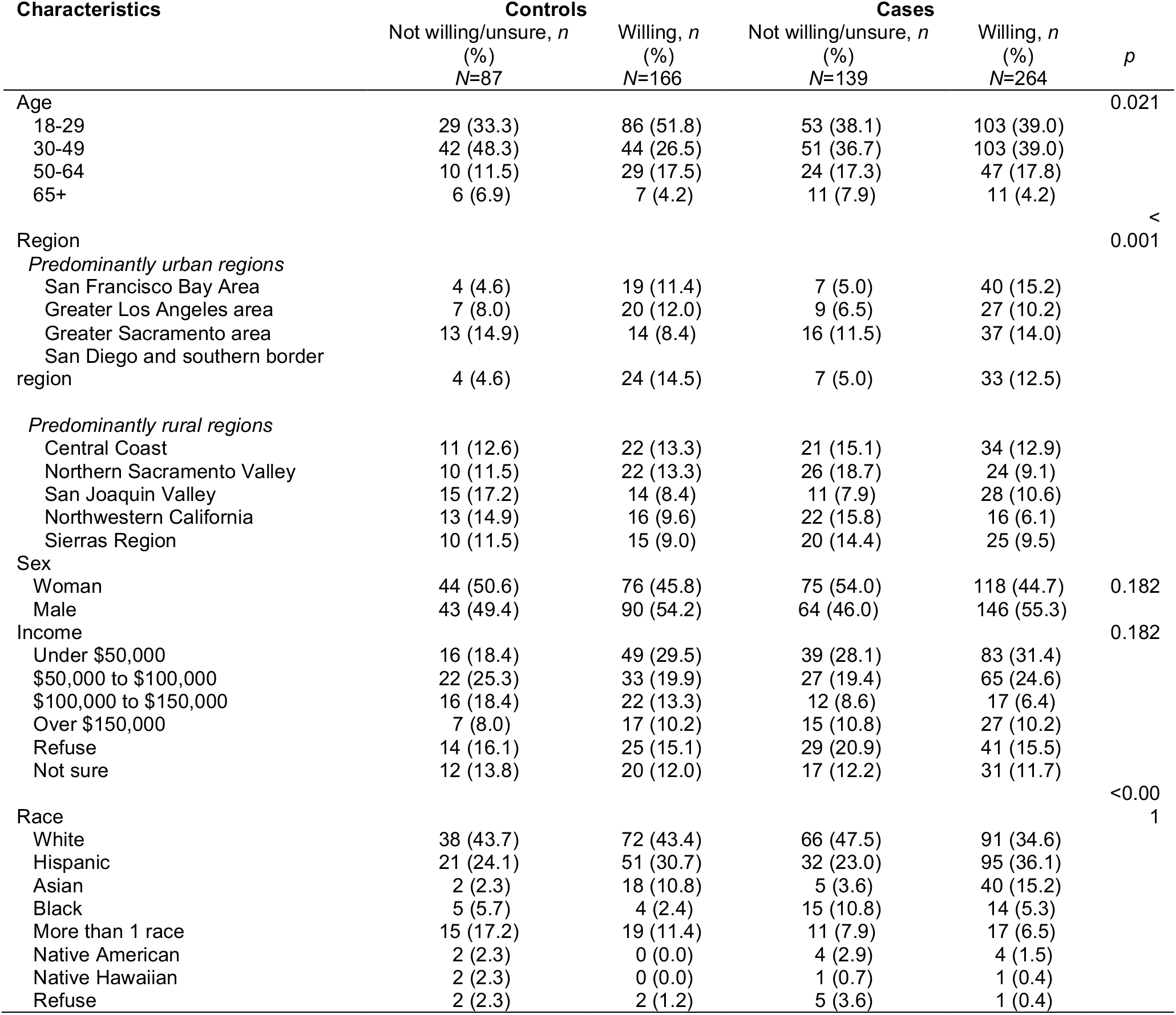
Vaccine confidence among cases and controls.

**Table S6:**
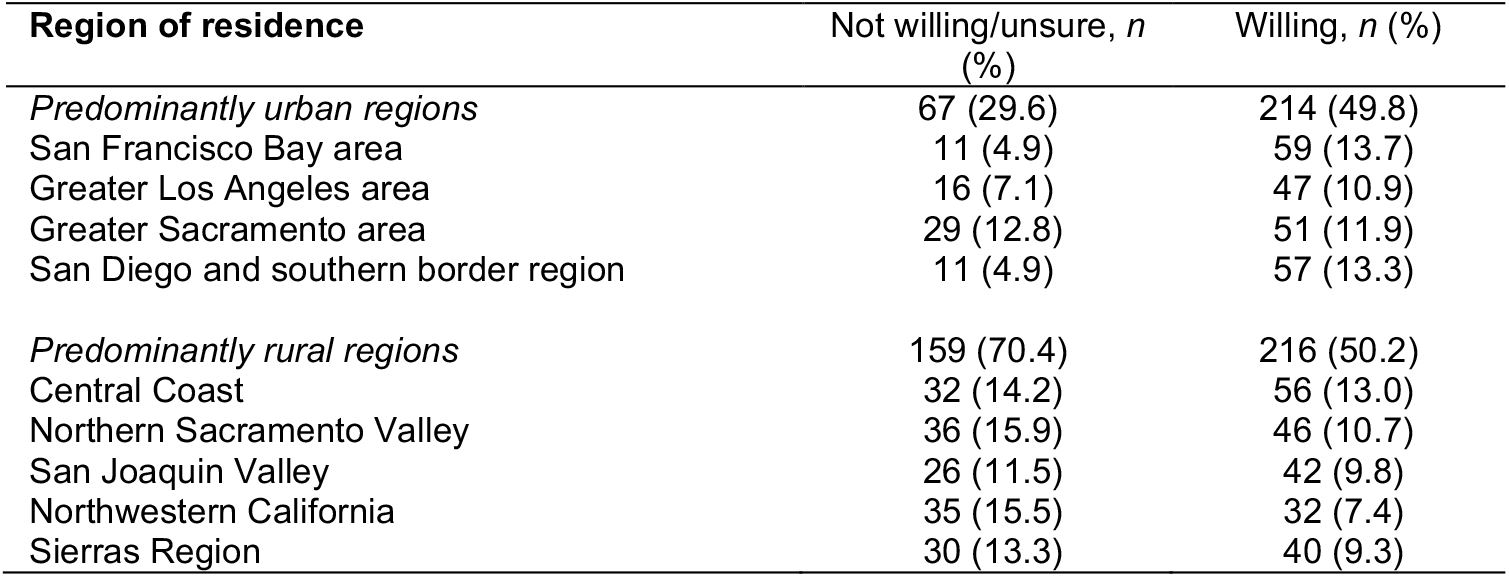
Regions of residence among participants reporting hesitancy or willingness to receive vaccination.

**Table S7:** Complete case analysis of predictors of vaccine confidence.

| Participant characteristics | Enthusiasm to receive vaccination |  | Adjusted Odds Ratio (95% CI) |
| --- | --- | --- | --- |
|  | Not willing/unsure, *n* (%)<br>*N*=226 | Willing, *n* (%)<br>*N*=430 |  |
| Case status |  |  |  |
| Case with SARS-CoV-2 infection | 139 (61.5) | 264 (61.4) | N/A |
| Uninfected control | 87 (38.5) | 166 (38.6) | N/A |
| Age |  |  |  |
| 18-29 | 82 (36.3) | 189 (44.0) | Ref. |
| 30-49 | 93 (41.2) | 147 (34.2) | 1.22 (0.25,2.37) |
| 50-64 | 34 (15.0) | 76 (17.7) | 1.26 (0.26,3.16) |
| 65+ | 17 (7.5) | 18 (4.2) | 1.42 (0.29,3.47) |
| Region |  |  |  |
| Predominantly urban regions | 67 (29.6) | 214 (49.8) | Ref. |
| Predominantly rural regions | 159 (70.4) | 216 (50.2) | 1.21 (0.23,8.54) |
| Sex |  |  |  |
| Male | 107 (47.3) | 236 (54.9) | Ref. |
| Woman | 119 (52.7) | 194 (45.1) | 1.10 (0.26,8.2) |
| Income<sup>4</sup> |  |  |  |
| Under \$50,000 | 55 (24.3) | 132 (30.7) | Ref. |
| \$50,000 to \$100,000 | 49 (21.7) | 98 (22.8) | 1.31 (0.20,3.24) |
| \$100,000 to \$150,000 | 28 (12.4) | 39 (9.1) | 1.63 (0.16,20.67) |
| Over \$150,000 | 22 (9.7) | 44 (10.2) | 1.69 (0.11,7.53) |
| Race<sup>5</sup> |  |  |  |
| White | 104 (46.0) | 163 (38.0) | Ref. |
| Hispanic | 53 (23.5) | 146 (34.0) | 1.44 (0.24,11.0) |
| Asian | 7 (3.1) | 58 (13.5) | 1.45 (0.27,7.08) |
| Black | 20 (8.8) | 18 (4.2) | 1.28 (0.27,10.65) |
| More than 1 race | 26 (11.5) | 36 (8.4) | 1.33 (0.14,3.61) |
| Native American or Alaskan Native | 6 (2.7) | 4 (0.9) | 1.21 (0.27,3.14) |
| Native Hawaiian or Pacific Islander | 3 (1.3) | 1 (0.2) | 1.36 (0.11,4.67) |

**Figure S1:**
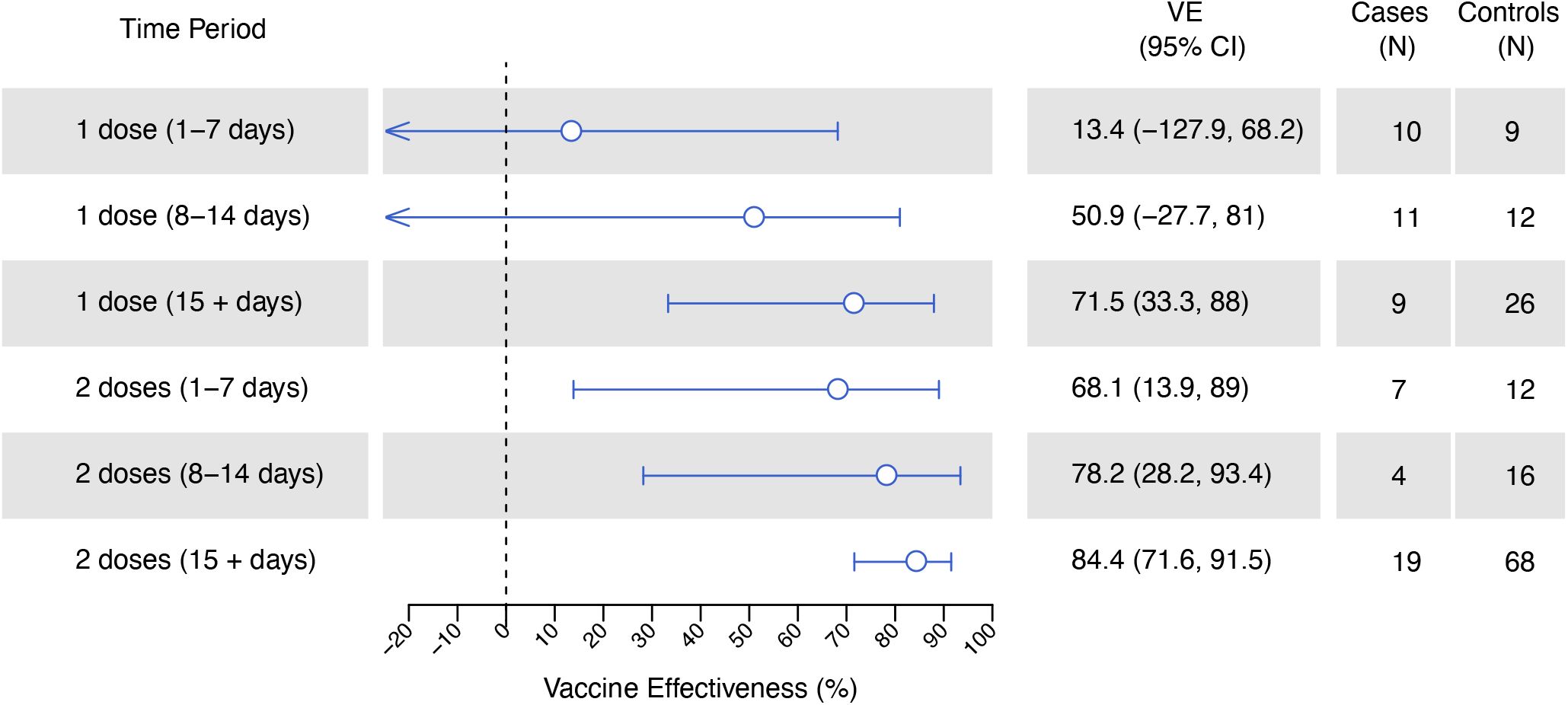
Sensitivity analyses of individuals (*N*=53) without access to vaccination cards. Lines denote 95% confidence intervals, respectively, for estimates of vaccine effectiveness for both mRNA vaccines poled. Estimates were calculated via conditional logistic regression.

## Notes

### Competing Interest Statement

JAL discloses receipt of grants and honoraria from Pfizer, Inc. unrelated to this work.

## REFERENCES

1. Polack FP, Thomas SJ, Kitchin N, et al. Safety and Efficacy of the BNT162b2 mRNA Covid-19 Vaccine. New England Journal of Medicine 2020; 383:2603–2615.

2. Baden LR, El Sahly HM, Essink B, et al. Efficacy and Safety of the mRNA-1273 SARS-CoV-2 Vaccine. New England Journal of Medicine 2021; 384:403–416.

3. Sadoff J, Le Gars M, Shukarev G, et al. Interim Results of a Phase 1–2a Trial of Ad26.COV2.S Covid-19 Vaccine. N Engl J Med 2021; :NEJMoa2034201.

4. State of California. Tracking COVID-19 in California - Coronavirus COVID-19 response. Available at: Covid19CaGov.

5. Patel MM, Jackson ML, Ferdinands J. Postlicensure Evaluation of COVID-19 Vaccines. JAMA 2020; 324:1939.

6. Lewnard JA, Patel MM, Jewell NP, et al. Theoretical framework for retrospective studies of the effectiveness of SARS-CoV-2 vaccines. Epidemiology, 2021. Available at: http://medrxiv.org/lookup/doi/10.1101/2021.01.21.21250258. Accessed 1 May 2021.

7. Amit S, Regev-Yochay G, Afek A, Kreiss Y, Leshem E. Early rate reductions of SARS-CoV-2 infection and COVID-19 in BNT162b2 vaccine recipients. The Lancet 2021; 397:875–877.

8. Thompson MG, Burgess JL, Naleway AL, et al. Interim Estimates of Vaccine Effectiveness of BNT162b2 and mRNA-1273 COVID-19 Vaccines in Preventing SARS-CoV-2 Infection Among Health Care Personnel, First Responders, and Other Essential and Frontline Workers — Eight U.S. Locations, December 2020–March 2021. MMWR Morb Mortal Wkly Rep 2021; 70:495–500.

9. Hall VJ, Foulkes S, Saei A, et al. COVID-19 vaccine coverage in health-care workers in England and effectiveness of BNT162b2 mRNA vaccine against infection (SIREN): a prospective, multicentre, cohort study. The Lancet 2021; :S014067362100790X.

10. State of California. Tracking Variants. 2021. Available at: https://www.cdph.ca.gov/Programs/CID/DCDC/Pages/COVID-19/COVID-Variants.aspx. Accessed 11 May 2021.

11. Honaker J, King G, Blackwell M. Amelia II: A Program for Missing Data. J Stat Soft 2011; 45. Available at: http://www.jstatsoft.org/v45/i07/. Accessed 2 September 2020.

12. Tenforde MW. Effectiveness of Pfizer-BioNTech and Moderna Vaccines Against COVID-19 Among Hospitalized Adults Aged ≥65 Years — United States, January–March 2021. MMWR Morb Mortal Wkly Rep 2021; 70. Available at: https://www.cdc.gov/mmwr/volumes/70/wr/mm7018e1.htm. Accessed 1 May 2021.

13. Reiter PL, Pennell ML, Katz ML. Acceptability of a COVID-19 vaccine among adults in the United States: How many people would get vaccinated? Vaccine 2020; 38:6500–6507.

14. Khubchandani J, Sharma S, Price JH, Wiblishauser MJ, Sharma M, Webb FJ. COVID-19 Vaccination Hesitancy in the United States: A Rapid National Assessment. J Community Health 2021; 46:270–277.

15. Fridman A, Gershon R, Gneezy A. COVID-19 and vaccine hesitancy: A longitudinal study. PLOS ONE 2021; 16:e0250123.

16. McCabe SD, Hammershaimb EA, Cheng D, et al. Unraveling Attributes of COVID-19 Vaccine Hesitancy in the U.S.: A Large Nationwide Study. medRxiv 2021; :2021.04.05.21254918.

17. Momplaisir F, Haynes N, Nkwihoreze H, Nelson M, Werner RM, Jemmott J. Understanding Drivers of Coronavirus Disease 2019 Vaccine Hesitancy Among Blacks. Clinical Infectious Diseases 2021; Available at: https://doi.org/10.1093/cid/ciab102. Accessed 1 May 2021.

18. Paterson P, Meurice F, Stanberry LR, Glismann S, Rosenthal SL, Larson HJ. Vaccine hesitancy and healthcare providers. Vaccine 2016; 34:6700–6706.

19. Grumbach K, Judson T, Desai M, et al. Association of Race/Ethnicity With Likeliness of COVID-19 Vaccine Uptake Among Health Workers and the General Population in the San Francisco Bay Area. JAMA Intern Med 2021; Available at: https://jamanetwork.com/journals/jamainternalmedicine/fullarticle/2778176. Accessed 1 May 2021.

20. Meyer MN, Gjorgjieva T, Rosica D. Trends in Health Care Worker Intentions to Receive a COVID-19 Vaccine and Reasons for Hesitancy. JAMA Netw Open 2021; 4:e215344.

21. Dagan N, Barda N, Kepten E, et al. BNT162b2 mRNA Covid-19 Vaccine in a Nationwide Mass Vaccination Setting. N Engl J Med 2021; 384:1412–1423.

22. Lewnard JA, Tedijanto C, Cowling BJ, Lipsitch M. Measurement of Vaccine Direct Effects Under the Test-Negative Design. Am J Epidemiol 2018; 187:2686–2697.

